# Expression of ACE2 receptor, soluble ACE2, Angiotensin I, Angiotensin II and Angiotensin (1-7), is modulated in COVID-19 patients

**DOI:** 10.1101/2021.02.08.21251001

**Authors:** Ikram Omar Osman, Cléa Melenotte, Philippe Brouqui, Matthieu Million, Jean-Christophe Lagier, Philippe Parola, Andréas Stein, Bernard La Scola, Line Meddeb, Jean-Louis Mege, Didier Raoult, Christian A. DEvaux

## Abstract

Although SARS-CoV-2 is primarily a pulmonary-tropic virus, it is nonetheless responsible for multi-organ failure in patients with severe forms of COVID-19, particularly those with hypertension or cardiovascular disease. Infection requires virus binding to the angiotensin I converting enzyme 2 (ACE2) monocarboxypeptidase, a regulator of blood pressure homeostasis through its ability to catalyze the proteolysis of Angiotensin II (AngII) into Ang(1-7). Although assumed, it had not been proven so far whether the SARS-CoV-2 replication in COVID-19 patients could modulate the expression of the ACE2 receptor and/or the AngII plasma levels. We demonstrate here, that in COVID-19 patients the ACE2 mRNA expression is markedly reduced in circulating blood cells. This ACE2 gene dysregulation mainly affects the monocytes which also show a lower expression of membrane ACE2 protein. Moreover, a significant decrease in soluble ACE2 plasma levels is observed in COVID-19 patients, whereas the concentration of sACE2 returns to normal levels in patients recovered from COVID-19. In the plasma of COVID-19 patients, we also found an increase in AngI and AngII. On the other hand, the plasma levels of Ang(1-7) remains almost stable in COVID-19 patients. Despite the Ang(1-7) presence in the plasma of COVID-19 patients it seems insufficient to prevent the effects of massive AngII accumulation. These are the first direct evidence that the SARS-CoV-2 may affect the expression of blood pressure regulators with possible harmful consequences on COVID-19 outcome.

## Introduction

The <u>S</u>evere <u>A</u>cute <u>R</u>espiratory <u>S</u>yndrome <u>Co</u>rona<u>V</u>irus-2, SARS-CoV-2, is the etiological agent of <u>CO</u>rona<u>VI</u>rus <u>D</u>isease (COVID)-19. Similarly to SARS-CoV that emerged in Asia in 2003, the SARS-CoV-2 that emerged at the end of 2019 is a *Betacoronavirus* lineage 2b/*Sarbecovirus* circulating in wildlife to which humans were susceptible(1,2). These two viruses shared 79.5% nucleotide identity(3-6). In recent months, the SARS-CoV-2 has spread worldwide (6-8). The virus can be responsible for severe COVID-19 forms characterized by multiple organ dysfunction syndrome (MODS) (9,10) as well as acute respiratory distress syndrome (ARDS), with high fatality risk (11). In 9 months, the COVID-19 pandemic caused more than 1 million deaths and 35 million confirmed cases worldwide according to the World Health Organization (WHO statistics, 5 October 2020).

Venous thromboembolism is a relatively common side effect of SARS-CoV-2 infection (up to one-third of critical COVID-19 cases), characterized by acute pulmonary embolism or intravascular coagulopathy that predisposes the patients to thrombotic events (12-14). Elevated D-dimers upon admission of patients is a biomarker of pulmonary embolism and is associated with increased mortality (15). Cytokines (such as IL-6) production is increased in severe COVID-19 and contributes in the “cytokine storm” leading to obstructive thrombo-inflammatory syndrome (16-18). Anticoagulants are considered a possible therapy to reduce the harmful circle of inflammation-coagulation-fibrosis observed in severe COVID-19 (19,20). To further understand the physiopathology of COVID-19, it is necessary to focus research on the viral receptor, the ACE2 molecule, and its function. The spike proteins (S1) from both SARS-CoV and SARS-CoV-2 were found able to bind the ACE2 expressed at the surface of human alveolar pneumocytes (21-23). Cell surface expression of ACE2 is however not limited to pneumocytes as the *ACE2* gene is differentially expressed in human tissues, offering a broad spectrum of cellular targets to the virus including enterocytes and the arterial and venous endothelial cells (24-26).

The interaction between S1 and ACE2 does not solely allow viruses attachment to target cells, it could also disturb the function of ACE2. Indeed, ACE2, a 805 amino acids type I cell-surface zinc-dependent monocarboxypeptidase, catalyzes the hydrolysis of the active vasoconstrictor octapeptide Ang II [Asp-Arg-Val-Tyr-Ile-His-Pro-Phe] to the heptapeptide Ang(1-7) and free L-Phe, thus controlling blood pressure (27-30). Therefore, ACE2 antagonizes the vasoconstrictor and profibrotic effects of AngII both by reducing the synthesis of AngII and by catalyzing the transformation of AngII into Ang(1-7) (31). ACE2 can also exert its functions through cleavage of AngI (the precursor of AngII) into Ang(1-9), which can get further metabolized into Ang(1-7) (32). Understanding the angiotensin pathway dysregulation in SARS-CoV-2 infected patients, could explain why one of the main comorbidities related to COVID-19 is hypertension (HT) (33,34), and why the administration of human recombinant soluble ACE2 (hrsACE2) to severe COVID-19 patients quickly improved their condition (35).

It has previously been reported that SARS-CoV, which is genetically very similar to SARS-CoV-2, modulates the expression of ACE2 (36-38). We have recently hypothesized (39) that a virus-induced decrease in ACE2 expression and/or inhibition of ACE2 peptidase function could result in an amplification loop leading to increased production of AngII expected to aggravate the severity of COVID-19 at the vascular level. To investigate this hypothesis we studied the expression of ACE2 mRNA, ACE2 cell-surface protein, as well as the plasma levels of sACE2, AngI, AngII, and Ang(1-7), in 44 patients (30 patients with COVID-19 disease and 14 post-COVID-19 patients) compared to 15 healthy volunteers. We demonstrate here that some of these biomarkers are significantly affected during COVID-19 and that these changes are conducive to worsening the patient’s condition.

## Materials and Methods

### Study Population

This study was coordinated by the IHU Méditerranée-Infection in Marseille (research protocol N° National:2020-A00864-35; ethical approval CPP 20.04.09/N°:20.04.01.83219/N°7626), and included 44 patients with a confirmed diagnosis of COVID-19 (these patients have all tested positive for SARS-CoV-2 in their nasopharyngeal samples as evidenced by a polymerase chain reaction (PCR) which uses specific primers for this virus. They were all subjected to a clinical examination including a pulmonary examination by low dose scanner. They all received hydroxychloroquine and azithromycin medication). The blood pressure whose values are known for certain patients (mainly those hospitalized at the IHU Méditerranée Infection), could not be reported for all patients (mainly those who came for routine outpatient visit). A follow up of the viral load was performed to monitor the effectiveness of treatment and the course of the disease at day 0, 3, 6 and 10. All healthy volunteers were tested negative for SARS-CoV-2 by PCR.

### Ethical Approval

This research protocol (N° National:2020-A00864-35) was submitted to ethical review by the French Participants Protection Committee (CPP East III; CPP president: Dr. P. Peton) prior to commencement of the study and received ethical approval CPP 20.04.09/N°:20.04.01.83219/N°7626. (Principal investigator, C. Devaux, Research Director at CNRS; Assay promotor : IHU Méditerranée Infection: Prof Didier Raoult, Director). An informed written consent was obtained from each participant to the protocol (both patients and healthy volunteers).

### Plasma collection

5 to 10 mL of blood samples was collected from patients and healthy donors into an Ethylenediaminetetraacetate (EDTA) tube (Sigma Aldrich, Saint-Quentin Fallavier, France), and allowed to clot at room temperature. The plasma was collected, then centrifuged at 1500g for 10 min and stored at −20 °C until use.

### PBMCs and adherent cells (monocytes) isolation

Peripheral blood mononuclear cells (PBMCs) were recovered from blood samples using Ficoll (Eurobio, Les Ulis, France) density gradient centrifugation followed by harvesting of PBMCs. Monocytes were isolated by adherence to culture dishes for 2 hours at 37°C and directly lysed for qRT-PCR without being harvested.

### Ribonucleic Acid (RNA) Extraction and Quantitative-Reverse Transcription Polymerase Chain Reaction (qRT-PCR)

RNAs were extracted from whole blood using PAXgene blood RNA kit (QIAGEN SA) with a DNase I step to eliminate DNA contaminants, according to the manufacturer instructions. The quantity and quality of the RNA was evaluated using a Nanodrop 1000 spectrophotometer (Thermo Science). The first-strand cDNA was obtained using oligo(dT) primers and Moloney murine leukemia virus-reverse transcriptase (MMLV-RT kit; Life Technologies), using 200 ng of purified RNA. The amplification cycles were performed using a real time PCR Mastercycler gradient (Eppendorf, Montesson, France). The qPCR experiments were performed using specific oligonucleotide primers (Specific primers were designed using the Primer3 software) and hot-start polymerase (SYBR Green Fast Master Mix; Roche Diagnostics). The specific primers used were Angiotensin Convertase Enzyme (ACE2) primers (forward primer: CAGGGAACAGGTAGAGGACATT and reverse primer: CAGAGGGTGAACATACAGTTGG) and the housekeeping β-Actin primers (forward: AGGAAGGAAGGCTGGAAGAG and reverse: GGAAATCGTGCGTGACATTA). The amplification cycles were performed using a C1000 Touch Thermal cycler (Biorad, Marnes-La-Coquette, France). The results of qRT-PCR were normalized using the housekeeping gene β-Actin (ACTB) and expressed as Relative Quantity (RQ= 2^-ΔCT^), where ΔCt = (Ct_Target−_ Ct_Actin_). The threshold cycle (Ct) was defined as the number of cycles required to detect the fluorescent signal.

### Quantification of plasma solubleACE2, angiotensin I, angiotensin II, and angiotensin1-7

The quantity of soluble ACE2, Angiotensin I, Angiotensin II; and Angiotensin 1-7 in the plasma of healthy volunteers and COVID-19 patients was determined using Elisa kits (FineTest, Wuhan, China). The experiments were performed according to the manufacturer’s instructions. The minimal detectable concentration of human soluble ACE2, Angiotensin I, Angiotensin II, Angiotensin (1-7) were 391 pg/mL, 125 pg/mL pg/mL, 31.2 pg/mL, and 15.6 pg/mL, respectively. The concentrations in each molecule were calculated by comparison to standard curves.

### Flow Cytometry

Flow cytometry was used to study the membrane expression of ACE2 as well as specific biomarkers on PBMCs. Cells were analyzed according to fluorescence intensity using an anti-ACE2-monoclonal antibody (mAb) labeled with Alexa Fluor 488 (R&D Systems, Minneapolis, USA) and mAb anti-CD3-PC5, anti-CD20-PC7, anti-CD16-PE, and anti-CD14-APC purchased from Beckman (Beckman coulter, Villepinte, France). Fluorescence intensity was measured using a Canto II cytofluorometer (Becton Dickinson, Biosciences, Le Pont de Claix, France) and the results were analyzed using a BD FACSDiva software v.6. 1.3 (Becton Dickinson, New Jersey, USA).

### Statistical Analysis and correlation analysis

Data were analyzed using a non-parametric Mann-Whitney U test to compare healthy donors, patients with COVID-19 and healed COVID-19 individuals. The results are presented as the median or the mean± SD. A p value <0.05 was considered statistically significant. The data were submitted to multivariate principal component analysis (PCA) biplot (score plot + loading plot) (RStudio) and hierarchical clustering heatmap analysis (ClustVis). The R studio program (RStudio, USA) was used to generate the correlation matrix for the biomarkers of interest after a Pearson test that measuring the linear correlation for two variables.

## Results

### Characteristics of patients

The clinical characteristics of the patients included in this protocol are summarized in **Table I**. None of the included patient had any severe or critical disease according to the WHO classification (40), only 4 patients required short hospitalization and none had to be placed in intensive care. All patients recovered from COVID-19 infection. Patients were divided into two groups. The first group, hereafter named COVID-19 patients (patients with viral persistence), was composed of 30 patients having difficulties to clear the virus and who remained positive for SARS-CoV-2 after 10 days therapy with hydroxychloroquine and 5 days azithromycine, including 5 men and 25 women whose mean age was 49.8 and 46.2 years, respectively. The second group, hereafter named healed COVID-19 patients (patients with rapid viral clearance), included 14 patients who had a marked decrease in their viral load at day 6 after therapy, including 7 men with a mean age of 48.6 years and 7 women with a mean age of 33.4 years. Samples were collected from patients and were then divided into the predefined two groups. Most of the patient samples used in this study were collected between 1 and 3 days after the treatment had been stopped (with exception of samples from IHU_14 and IHU_15 patients collected 5 days after the end of treatment, and samples from IHU_17 and G2 patients collected respectively 7 days and 10 days after the end of treatment). A group of 15 healthy volunteers was included in this study as control among which 7 males mean age 40.3 years and 8 females mean age 26.9 years. Samplings were performed between 13 and 14 days after COVID-19 diagnosis (i.e. less than 5 days after the test turned negative on the nasopharyngeal sample for the COVID-19 patient group and more than 5 days after qRT-PCR turned negative on nasopharyngeal sample for the healed patients group.

**Table I:**
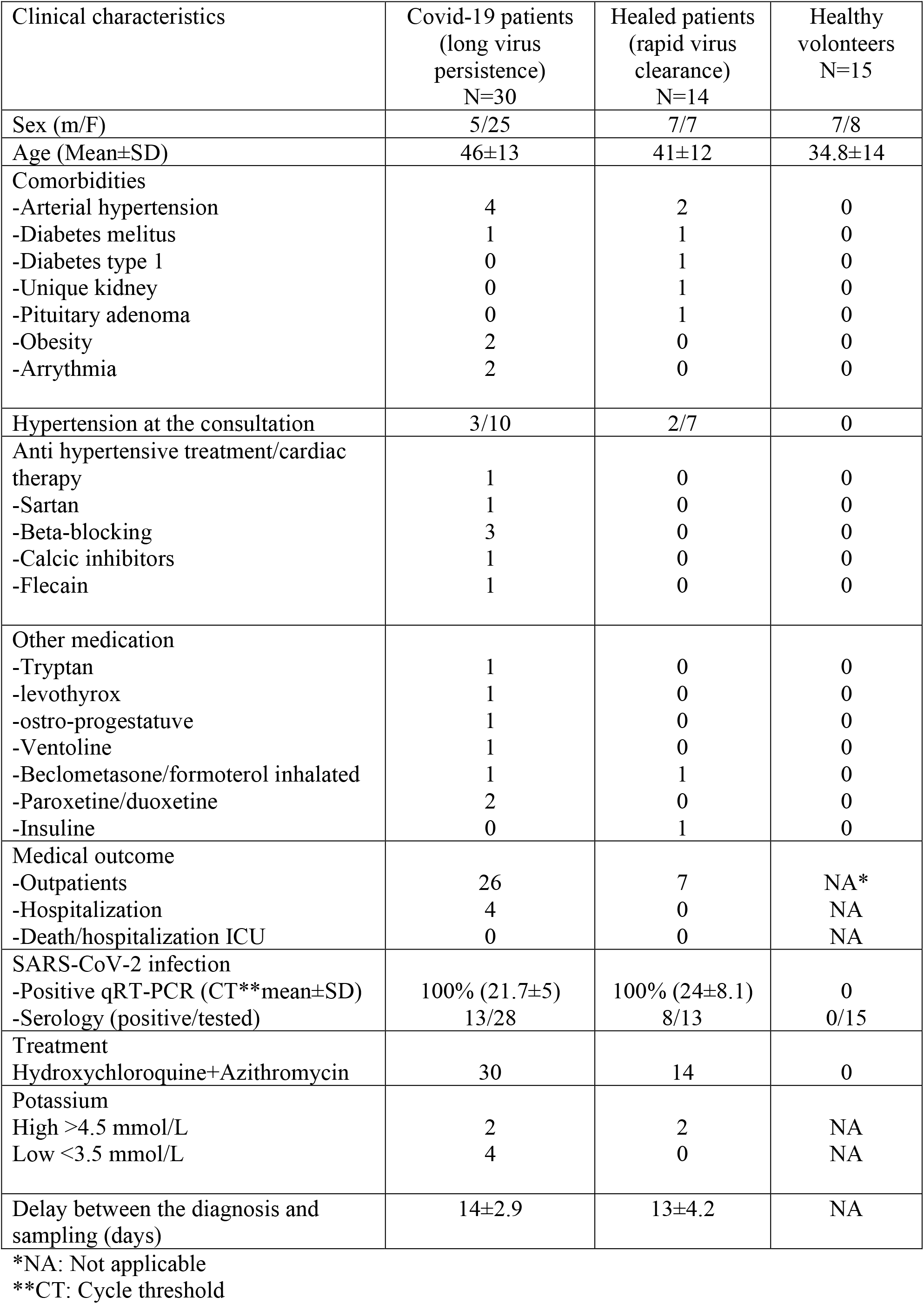
Patients/clinical data.

| Clinical characteristics | Covid-19 patients<br>(long virus<br>persistence)<br>N=30 | Healed patients<br>(rapid virus<br>clearance)<br>N=14 | Healthy<br>volunteers<br>N=15 |
| --- | --- | --- | --- |
| Sex (m/F) | 5/25 | 7/7 | 7/8 |
| Age (Mean±SD) | 46±13 | 41±12 | 34.8±14 |
| Comorbidities |  |  |  |
| -Arterial hypertension | 4 | 2 | 0 |
| -Diabetes melitus | 1 | 1 | 0 |
| -Diabetes type 1 | 0 | 1 | 0 |
| -Unique kidney | 0 | 1 | 0 |
| -Pituitary adenoma | 0 | 1 | 0 |
| -Obesity | 2 | 0 | 0 |
| -Arrhythmia | 2 | 0 | 0 |
| Hypertension at the consultation | 3/10 | 2/7 | 0 |
| Anti hypertensive treatment/cardiac<br>therapy | 1 | 0 | 0 |
| -Sartan | 1 | 0 | 0 |
| -Beta-blocking | 3 | 0 | 0 |
| -Calcic inhibitors | 1 | 0 | 0 |
| -Flecain | 1 | 0 | 0 |
| Other medication |  |  |  |
| -Tryptan | 1 | 0 | 0 |
| -levothyrox | 1 | 0 | 0 |
| -ostro-progestative | 1 | 0 | 0 |
| -Ventoline | 1 | 0 | 0 |
| -Beclometasone/formoterol inhaled | 1 | 1 | 0 |
| -Paroxetine/duoxetine | 2 | 0 | 0 |
| -Insuline | 0 | 1 | 0 |
| Medical outcome |  |  |  |
| -Outpatients | 26 | 7 | NA* |
| -Hospitalization | 4 | 0 | NA |
| -Death/hospitalization ICU | 0 | 0 | NA |
| SARS-CoV-2 infection |  |  |  |
| -Positive qRT-PCR (CT**mean±SD) | 100% (21.7±5) | 100% (24±8.1) | 0 |
| -Serology (positive/tested) | 13/28 | 8/13 | 0/15 |
| Treatment |  |  |  |
| Hydroxychloroquine+Azithromycin | 30 | 14 | 0 |
| Potassium |  |  |  |
| High >4.5 mmol/L | 2 | 2 | NA |
| Low <3.5 mmol/L | 4 | 0 | NA |
| Delay between the diagnosis and<br>sampling (days) | 14±2.9 | 13±4.2 | NA |

\*NA: Not applicable
\*\*CT: Cycle threshold

### Reduced ACE2 mRNA expression in circulating blood cells from COVID-19 patients

In order to assess the effect of SARS-CoV-2 on the *ACE2* gene expression in people hospitalized or having consulted for symptoms related to COVID-19 disease, we used qRT-PCR to compare the ACE2 mRNA expression in circulating cells from 30 COVID-19 disease patients and 14 patients healed from COVID-19 with 15 healthy volunteers (**Figure 1A**). When the median values of ACE2 mRNA expression were compared among groups, a reduced expression of ACE2 mRNA was observed in COVID-19 patients compared to healthy volunteers (median values 0.722 x 10^−3^ versus 1.433 x 10^−3^). Yet, this result appeared not statistically significant according to Mann-Whitney test (p=0.0577). This tendency to downregulation of ACE2 gene expression was also observed in people healed from COVID-19 disease compared to healthy volunteers (median values 0.540 x 10^−3^ versus 1.433 x 10^−3^), and even accentuated compared to the COVID-19 group. The reduced expression of ACE2 mRNA in patients healed from COVID-19 was statistically significant according to Mann-Whitney test (p=0.0307). In addition, there were no significant differences between the two groups of patients (p=0.2771). Moreover, treatment for hypertension (HT) appears to promote a more rapid return to normal *ACE2* gene expression since, in the healed group, patients under anti-HT treatment have the closest level of ACE2 expression to the average levels of ACE2 mRNA expression observed in healthy volunteers. We confirmed these results by investigating the expression of ACE2 mRNA in PBMCs, instead of the whole blood circulating cells (the difference resides in the lack polynuclear cells in PBMCs) (**Figure 1B**). Out of the 44 people from the patients’ groups, 28 patients were tested irrespective of the level of ACE2 mRNA expression in their PBMCs. Under these new experimental conditions, the results were very similar to the previous ones with a tendency to reduced ACE2 mRNA expression in the two groups of patients who were infected by SARS-CoV-2 compared to healthy volunteers (median values 0.561 x 10^−3^ and 1.28 x 10^−3^ versus 2.18 x 10^−3^ for COVID-19 patients, patients healed from COVID-19 and healthy volunteers, respectively). This downregulation of ACE2 gene expression in the COVID-19 patients’ group (n=10), was found significant compared to the group (n=8) of healthy volunteers (p=0.0343), according to Mann-Whitney test. Although a comparison of the median values of recovered COVID-19 patients with those in the healthy volunteers group (median values 1.28 x 10^−3^ versus 2.18 x 10^−3^), showed a down-regulation trend in ACE2, this result is not statistically significant (p=0.0752).

**Figure 1:**
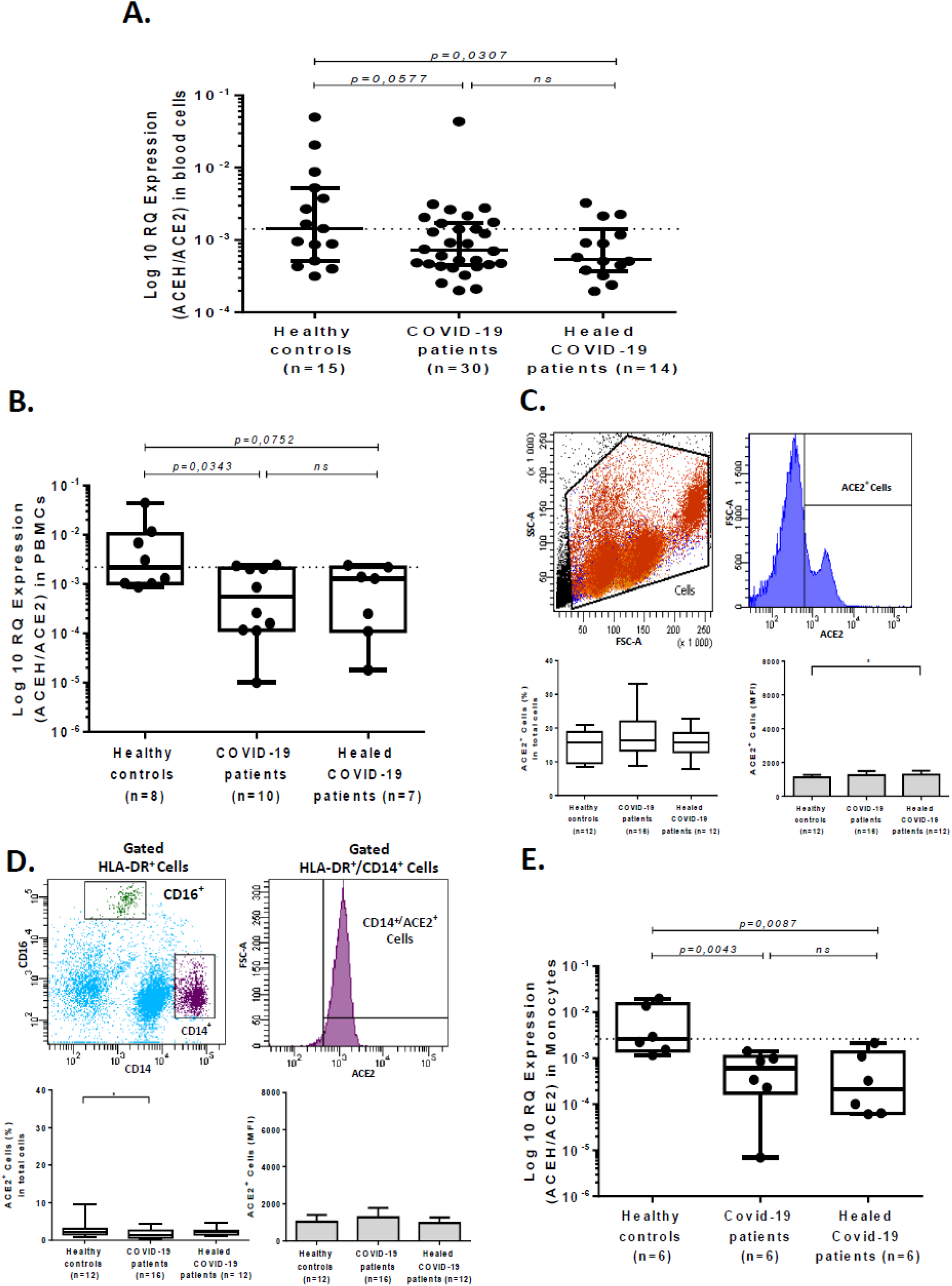
ACE2 mRNA expression in blood cells, PBMCs, and monocytes from COVID-19 patients and healthy volunteers. A. Expression of ACE2 mRNA in circulating blood cells (both mononuclear and polynuclear cells) from the group of patients with COVID-19 (n = 30) or patients healed from COVID-19 (n = 14) compared to a healthy volunteers control group (n= 15). The results are expressed as log10 RQ were RQ = 2^(-ΔCT)^. B. Expression of ACE2 mRNA in PBMCs samples from the three different groups studied: patients with COVID-19 (n = 8), patients healed from COVID-19 (n = 10), and healthy volunteers (n= 7). C. Flow cytometry analysis of ACE2 expression at the surface of PBMCs from patients with COVID-19 (n=16) and patients healed from COVID-19 (n=12) versus PBMCs from healthy volunteers (n=12). The upper panels show the gating parameter chosen (from individual to individual, between 8 and 35% of PBMCs express ACE2 at their surface), the lower left panel indicates the percent of cells expressing ACE2 while the lower right panel is an histogram representing the mean fluorescence intensity (MFI). D. Flow cytometry analysis of ACE2 protein expression at the surface of different cell populations during COVID-19. The gating (upper panels) was performed using different cluster differentiation-specific mAb (CD14, CD16, HLA-DR). The figure illustrates the results obtained with the HLA-DR^+^/CD14^+^ population while data obtained with respect to other cell populations are presented in the supplementary figure S1.The lower panels show the percent of monocytes expressing ACE2 (left) and their ACE2 fluorescence intensity (right). E. qRT-PCR analysis of ACE2 mRNAs expression in monocytes cells from COVID-19 patients (n = 6) and patients healed from COVID-19 (n = 6) versus monocytes cells from healthy donors (n = 6). The data are expressed as log10 RQ were RQ = 2^(-ΔCT)^. Mann– Whitney test used for the statistical analysis.

When considering circulating whole blood cells or PBMCs, in both cases the median values of ACE2 mRNA expression in patients were lower than those measured in healthy volunteers. These results indicate that the decrease in the ACE2 gene transcription can be considered a biomarker of COVID-19, as it was observed during the symptomatic phases and prolonged in people who had undetectable viral load and had recovered.

### Decreased ACE2 protein expression in monocytes from COVID-19 patients

Since ACE2 mRNA expression was found down-regulated in circulating blood cells from COVID-19 patients and patients healed from COVID-19, we investigated whether this could be reflected in the cell-surface expression of the ACE2 protein. To this end, after incubation with Alexa Fluor 488-labeled anti-ACE2 mAb, PBMCs from patients and healthy volunteers were examined by flow cytometry. We noticed a strong heterogeneity in PBMCs ACE2 surface expression among individuals within each group (between 8 and 35% of PBMCs expressed ACE2). ACE2 expression at the surface of PBMCs from patients with COVID-19 (n=16) and patients healed from COVID-19 (n=12), was quantified and compared to that of PBMCs from healthy volunteers (n=12) (**Figure 1C**). Despite the observed tendency to ACE2 mRNA dowregulation in PBMCs from patients (see Figure 1B), the percentage of PBMCs expressing membrane-bound ACE2 remained almost stable or slightly increased in the two groups of patients compared to healthy volunteers. To try to understand this apparent paradox, the expression of ACE2 on the surface of separated cell populations was studied. The modulation of ACE2 cell-surface expression differed from one cell population to another as illustrated by the significant increase in the number of CD3^+^ T cells expressing ACE2 in COVID-19 patients, while there was an absence of modulation in the CD20^+^ B cells and the CD16^+^/HLA-DR^+^ monocytic/dendritic cells expressing ACE2 compared to cell populations from healthy volunteers (**Figure S1 Supplementary data**). In addition, we observed (**Figure 1D**) a slight but significant decrease in the number of CD14^+^/HLADR^+^ monocytes that expressed membrane-bound ACE2 in the COVID-19 patients (n=16). To conclude whether or not the decrease in the expression of ACE2 on the monocytes that represent only a small percentage of PBMCs, could by itself account for the apparent decrease in ACE2 mRNA expression in the whole PBMCs and circulating blood cells, the ACE2 mRNA expression was quantified in adherent monocytic cells obtained from several patients and healthy volunteers (**Figure 1E**). A statistically significant reduction of ACE2 mRNA expression was found in adherent monocytic cells from COVID-19 patients (p=0.0043) and patients healed from COVID-19 (p=0.0087), compared to the same cell type in healthy volunteers.

Our results indicate that membrane-bound ACE2 expression is modulated among populations of circulating blood cells during COVID-19 and that there was a significant decrease in the expression of ACE2 mRNA and cell-surface ACE2 protein in the monocytes of patients who were infected with SARS-CoV-2.

### Reduced plasma levels of sACE2 in COVID-19 patients

The release of soluble ACE2 (sACE2) in plasma was investigated using an ELISA assay (**Figure 2A**). The expression of sACE2 was found heterogeneous among the healthy control group. Three patients (code: C11, C06, and C07) of the 15 healthy controls, were found with a sACE2 well above the other group members, among which a man in his 30’s characterized by a high expression of ACE2 mRNA. In the COVID-19 patients’ group, there was also a strong heterogeneity with 5 patients (code: IHU_5, IHU_8, IHU_42, IHU_21 and IHU_44), for whom the expression was much higher than for the 25 other members of the group. All 5 patients were women, including two with high blood pressure, but none of them with elevated expression of ACE2 mRNA. Regarding the healed group, the results were more homogeneous. Despite these individual variations, what was remarkable according to a Mann-Whitney test, was that the plasma level of sACE2 which was found to be significantly decreased in the COVID-19 group, compared to the healthy volunteers’ group (19396 pg/mL versus 22600 pg/mL; p=0.0156). It should also be noted that the healed COVID-19 group had a plasma level of sACE2 of 22141 pg/mL, which was not significantly different from the healthy control group (p=0.153).

**Figure 2:**
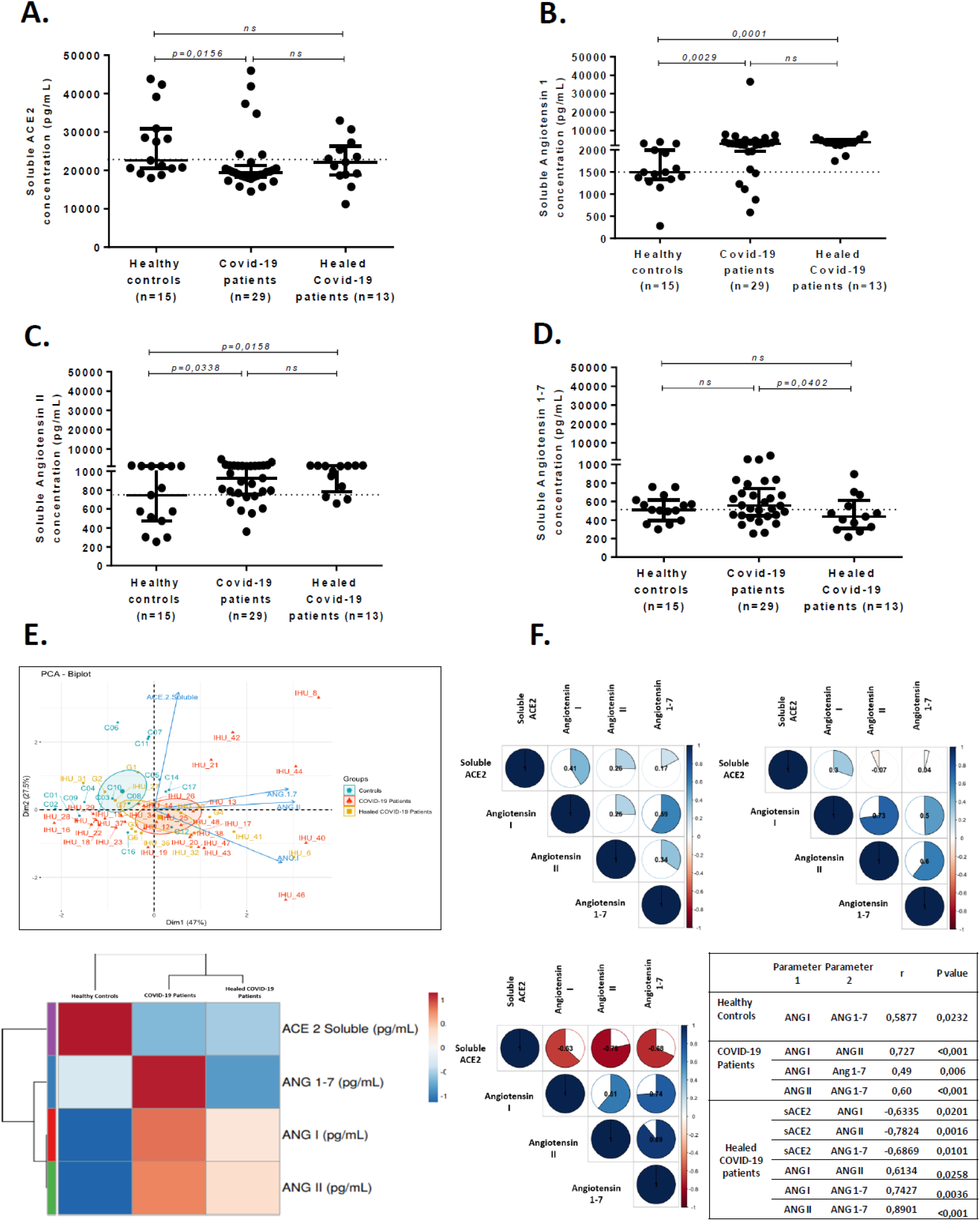
ELISA quantification of soluble sACE2 protein, and angiotensin metabolites Ang I, AngII, and Ang(1-7) in different groups of COVID-19 patients and healthy volunteers. Quantification of sACE2 (A), AngI (B), AngII (C), and Ang(1-7) (D) in the plasma of COVID-19 patients (n=29) and patients healed from COVID-19 (n=13) compared to the healthy volunteers control group (n=15). The Mann–Whitney test was used for the statistical analysis. E. The top panel represents the principal component analysis (PCA) biplot (score plot + loading plot), and the lower panel illustrates the hierarchical clustering heatmap analysis (each colored cell on the map corresponds to a concentration value) of the different angiotensin metabolites in each group. F. Correlation matrix (R studio used to generate the matrix). The upper panels show the correlation matrix of the different biomarkers of interest for the group of healthy volunteers (left) and COVID-19 patients (right), while the correlation matrix for the group of patients healed from COVID-19 is shown in the lower left panel. The table at the lower right part of the figure summarizes the correlation analyses (Pearson test and/or Spearman test) between the different metabolites (AngII, Ang(1-7) and sACE2 in each group.

These results strongly suggest that the significant reduction of sACE2 found in COVID-19 patients was a direct consequence of the SARS-CoV-2 infection and corroborated the reduction in expression of ACE2 mRNA and membrane-bound ACE2. Insofar as healed COVID-19 patients regain a normal level of serum sACE2 once patients had cleared their virus, this molecule could be considered a biomarker of the progression of the disease with a plasma decrease in patients with detectable viral load and a progressive return to normal in patients with rapid virus clearance.

### Increased plasma levels of Ang I and Ang II in COVID-19 patients

Since ACE2 is a key peptidase that regulates the renin-angiotensin-aldosterone system (RAAS), it was of major importance to investigate the plasma level of the main peptides of this pathway. The quantification of AngI, a peptide resulting from the cleavage of the angiotensinogen by renin was performed as it serves as upstream precursor for AngII production following hydrolysis by the ACE peptidase. In addition, since ACE2 expression was found to be reduced in patients with COVID-19, it is of interest to study whether this affected the plasma concentration of AngII. For this purpose, we used AngI and AngII ELISA kits. The comparison of AngI levels found in healthy volunteers (median=1496 pg/mL), with that from patients showed a significant increase in AngI in both COVID-19 patients (median= 2979 pg/mL; p=0.0029) and patients healed from COVID-19 (median= 3603 pg/mL; p=0.0001) (**Figure 2B**). Next, AngII was quantified in the patient’s plasma and that of the healthy volunteers. This study showed that the serum AngII concentration was higher in the COVID-19 patients (median= 921 pg/mL and 1038 pg/mL for COVID-19 patients and patients healed from COVID-19, respectively), than in healthy volunteers (median=746 pg/mL) (**Figure 2C**). The differences between the two groups of patients and the healthy volunteers were found to be significant according to a Mann-Whitney test (p=0,0190 and p=0,0099, respectively), and was also confirmed by the principal component analysis and hierarchical clustering heatmap which show that the two groups of patients behave differently from the control group for all parameters studied (**Figure 2E**). It is noteworthy that the plasma concentration of AngII was extremely high in two patients from the COVID-19 patients group. These patients were two women, respectively in their late-40’s (patient code: UHI_24) and mid-50’s (patient code: IHU_46), without documented history of high blood pressure and anti-HT treatment. Their kalemia values were also in the normal range (4.06 mmol/L and 4.1 mmol/L, respectively).

These data indicate that, in COVID-19 patients, a reduced production of ACE2 synthesis is accompanied by an increase in AngII plasma levels. Since the RAAS pathway is also disturbed upstream to the AngII synthesis, it can be hypothesized that the accumulation of AngII is either linked to an over-production of AngI, a lower conversion rate of AngII into Ang (1-7) by ACE2, or both.

### Stable plasma levels of Ang(1-7) in COVID-19 patients

The finding that AngII accumulated in the plasma of COVID-19 patients prompted us to identify the plasma levels of Ang(1-7), the downstream molecule of the RAAS pathway. We studied the Ang(1-7) plasma concentrations in the 3 studied groups (**Figure 2D**). Indeed we found a slight increase in the Ang(1-7) plasma concentrations between healthy volunteers (median: 510 pg/mL), and COVID-19 patients (median: 554 pg/mL). We noted in particular a strong expression of Ang(1-7) in three COVID-19 patients (patients code: IHU_24, IHU_8 and IHU_13), the IHU_24 patient (see above) with the highest serum level of AngII in this series, the patient IHU_8, a old woman in her 80’s with HT (candesartan medication; kalemia: 3.68 mmol/L), while the patient IHU_13 was a old man in his 50’s with no known history of HT (kalemia: 3.82 mmol/L). In fact, according to the Pearson test (**Figure 2F**), it appears that the increase in AngI causes a simultaneous increase in Ang(1-7) (r=0.49, p <0.006 in COVID-19 patients and r=0.74, p=0.0036 in patients healed of COVID-19), which could suggest that the neprilysin/thimet oligopeptidase alternative pathway of Ang(1-7) biosynthesis from AngI is activated when the Ang(1-7) production through AngII cleavage by ACE2 is impaired. Regarding AngII, we found that the increase in AngII has also caused a simultaneous increase in Ang(1-7) and this effect was statistically significant (r=0.60, p <0.001 in COVID-19 patients and r=0.89, <0.001 in patients healed of COVID-19). Yet, as shown in figure 2D, we also found a statistically significant decrease in Ang(1-7) between COVID-19 patients (median: 554 pg/mL) and patients healed from COVID-19 (median: 436 pg/mL; p=0.00402). There is also a significant inverse correlation (r=-0.78; p= 0.0016) between decreased sACE2 and increased plasma AngII concentration in patients cured of COVID-19.

These data suggest that in COVID-19 patients there is no reduction of the plasma concentration of Ang(1-7) despite reduced ACE2 catalytic activity and that the abnormal high concentration of AngI serves as substrate for Ang(1-7) biosynthesis through the neprilysin/thimet oligopeptidase alternative pathway of Ang(1-7) biosynthesis. In parallel, the plasma concentration of AngII increase since there is a decreased metabolism of this molecule through the ACE2 peptidase.

## Discussion

During the past nine months, *in vitro* and *in silico* studies allowed to analyze interactions between SARS-CoV-2 and ACE2 (41-43). This has led many clinical research teams to hypothesize that soon after infection of people, SARS-CoV-2 is likely to trigger a dysregulation of ACE2 and subsequently AngII and Ang(1-7), contributing to worsening hypertension and releasing of proinflammatory cytokines, especially IL-6, thereby accelerating endothelial dysfunction and atherogenesis (34,44). ACE2 is thought to act in an opposing manner to its homologue, angiotensin-converting enzyme (ACE), by inactivating the vasoconstrictor peptide AngII and generating the vasodilatory fragment, Ang(1–7) (45). As illustrated by **Figure 3**, we report here the first direct *in vivo* evidence that ACE2 mRNA, ACE2 protein, sACE2, AngI, AngII and Ang(1-7) expression are modulated in COVID-19 patients.

In order to conduct this study, we only had access to blood samples, because it is a simple medical act, not particularly invasive, and it can be performed with the informed consent of the patient and the agreement of the ethics committee. However, one must keep in mind that it is mainly at the level of the pulmonary, vascular and cardiac epithelial tissues that the impact of the SARS-CoV-2 should be the greatest since it is at the level of epithelial tissues that ACE2 molecules are primarily expressed (46-48). Although the *ACE2* gene is usually considered silent in immune cells, the expression of ACE2 mRNAs was previously reported in a subset of CD14^+^ CD16^-^ human monocytes (49), opening to us the possibility of conducting a study using circulating blood cells. There is not such limitation for the quantification of soluble compounds (sACE2, AngI, AngII and Ang(1-7) in plasma. Regarding the cohort of 44 patients we studied, there is a possible bias due to their sex, age, and treatment. It is generally considered that among patients with symptomatic COVID-19, about two-thirds are men whereas in our studied group of COVID-19, men represented only 16.6% of the group. Another possible bias lies in the recruitment of patients with 83.4% women in the COVID-19 group to be compared to 46.6% in the healthy control group. Regarding the plasma concentration of sACE2, literature reports that males express little higher ACE2 than females (46, 50). This can be linked to the finding that ACE2 is encoded by a gene mapping to chromosomal position Xp22 (32), and is in agreement with the observation that conversion of Ang II to Ang (1-7) by ACE2 was higher in males than females (51). However, in the small number of cases studied, we found no significant difference between males and females for the expression of ACE2 mRNA. At the time of first clinical examination, although blood pressure was verified for all patients, it was only recorded on the admission sheet for 15/44 patients. In the COVID-19 patients group, 4 patients (patient code: IHU_12, IHU_21, IHU_40, and IHU_44) showed hypokalemia while 2 patients showed hyperkalemia (patient code: IHU_15 and IHU_48), and in the group of patients healed from COVID-19, 2 patients showed hyperkalemia (patient code: IHU_27, and IHU_30). In the two groups of patients, among 44 individuals, seven were known to have a history of hypertension and two more were found with high blood pressure at the time of clinical examination. Anti-HT therapy used by the patients were recorded and indicated in **Table I**. In addition, worldwide ACE (52-54) and ACE2 polymorphisms (55-57), are thought to change the AngII plasma levels and affect COVID-19 severity and recovery rate. It was previously claimed that Asian people express ACE2 higher than Caucasian and African American populations (46). In the IHU Méditerranée Infection cohort of patients, this cannot be evaluated because it is strictly forbidden to record the ethnic origin of treated patients, so we have no information on the genetic background of the patients studied, even though it is obvious that Marseille is a melting point for population mixing and that the ethnic origins of the patients are diverse. Finally, regarding the patients’ COVID-19 therapy, all these patients received 600 mg of hydroxychloroquine (HCQ) daily during 10 days (plus or minus 1 day) and azithromycin was added to their treatment during the first 5 days (this therapy is routinely used for COVID-19 patients admitted in our institute) (58). It cannot be excluded that the HCQ treatment could modified the ability of the virus to bind to the ACE2 receptor since among other *in vitro* antiviral effect of HCQ on SARS-CoV-2 it was suggested that a HCQ mediated deficit in the glycosylation of the ACE2 virus cell surface receptor might affect virus attachment (59, 60). Despite the high heterogeneity of ACE2 mRNA expression from individual to individual within each group, we observed a trend of lower ACE2 mRNA expression in both patient groups compared to the healthy volunteers. It is worth noting that the level of ACE2 Mrna expression in the healthy volunteers group was highly heterogeneous, particularly regarding three volunteers (patient code C10, C17, and C11, respectively), a male in his 60’S, a female in her late-20’s, and a 32-years-old male in his early-30’s, who had elevated basal ACE2 mRNA expression. The difference in ACE2 mRNA expression was significant between the healthy control group and the healed COVID-19 group (p=0.0307), as if the expression of ACE2 retained the imprint of the infection for some time after viral clearance and recovery. In the COVID-19 patient groups, one patient (patient code: IHU_7) with an ACE2 expression significantly higher than the mean value for this group was a obese woman in her late-40’s on analgesic therapy (nefopam). Her COVID-19 history was marked by a extended viral carriage during 21 days. In the group of patients healed from COVID-19, the three patients with the slightly higher ACE2 mRNA expression were male patients, the IHU_27 was a patient in his late-50’s with hypertension (196/85) and hyperkalemia (4.65 mmol/L), IHU_6 was a patient in his 60’s also with above-normal blood pressure (159/95) and a normal kalemia of 3.97 mmol/L, both patients were on anti-HT treatment, while the third one IHU_36 was male patients in his 30’s (kalemia: 3.91 mmol/L), treated for allergy and inflammatory bronchial obstruction. However, the median value of ACE2 mRNA expression in those patients groups remained below the median value of ACE2 mRNA expression in the healthy control group. It is generally accepted that *in vitro* binding to ACE2 and/or Sarbecovirus infection of cells reduces the expression of ACE2 mRNA (37). In the absence of Sarbecovirus infection, reduced levels of ACE2 expression were previously reported in cardiac tissues linked to HT, dyslipidemia and/or heart failure (61,62). The ACE2 mRNA down-regulation observed in COVID-19 patients and patients healed from COVID-19 could also be the result of a feedback regulation loop orchestrated by AngII. It was previously reported that the over-expression of AngII decreased ACE2 mRNA production in rat cardiac myocytes and fibroblasts. This ACE2 mRNA down-regulation was blocked by AT1 receptor blocker losartan and inhibitors of MAP kinases, suggesting that AngII triggers AT1 receptor signaling that leads to Erk1/Erk2 activation and modulation of ACE2 expression (63). In previous opinion papers, we and others (31,64-67) strongly encouraged to rapidly evaluate whether the Renin Angiotensin System (RAS) blockers (namely the ACE inhibitors, ACEi, and AngII receptor blockers, ARBs), aimed at treating HT, are more beneficial than harmful in severe COVID-19 patients. Here we have found preliminary evidence that being on HT appears to promote a faster return to normal ACE2 gene expression in cured COVID-19 patients. Although we had not enough patients with HT medication to draw definitive conclusion, this observation argues in favor of continuing RAS inhibitors therapy during COVID-19 disease. It corroborates recent data indicating that RAS blockers improve the clinical outcomes of COVID-19 patients with HT (68-70).

**Figure 3:**
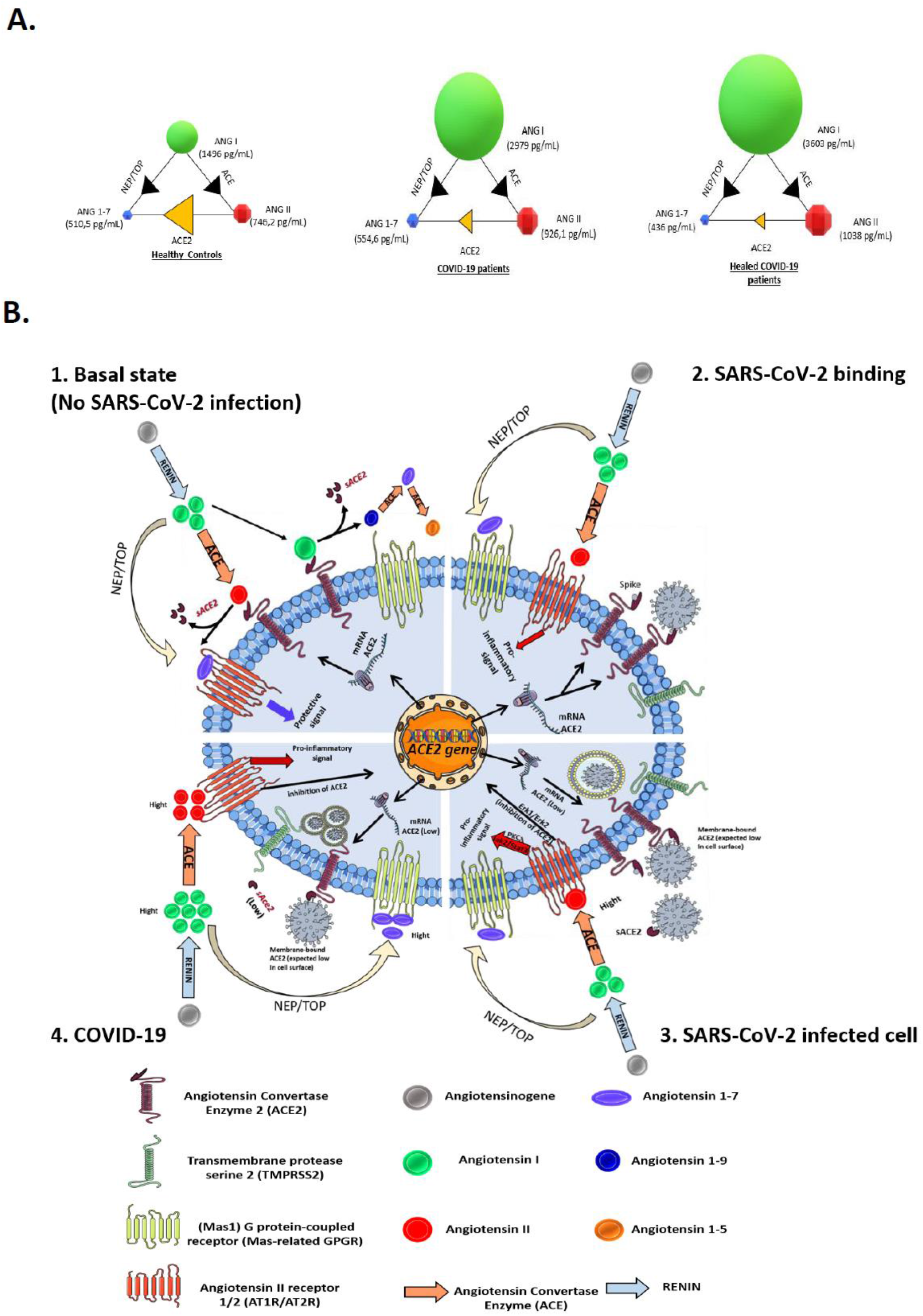
Schematic representation of the possible dysfunction of the angiotensin metabolites pathway in COVID-19 patients. A. Modelization of the concentration of angiotensin metabolites in plasma in healthy volunteers, COVID-19 patients, and patients healed from COVID-19. For each group, the individual circles indicate the angiotensin metabolite median concentrations, shown in pg/mL. The arrows between the circles mark the catalyzing reactions mediated by the indicated enzymes. B. Schematic diagram of the renin-angiotensin system (RAS) cascade in a normal physiological state and in COVID-19 patients. Renin cleaves Angiotensinogen to Ang I which is further processed to the vasoconstrictor AngII peptide by ACE. The upper left panel illustrates the AngII pathway of Ang (1-7) biosynthesis where ACE2 converts Ang II to Ang(1-7) and also AngI to Ang (1-9) next converted in Ang (1-7) by ACE peptidase, leading to protective signal through MAS-related GPGR. This is the physiological state. The upper-right panel illustrates the possible dysfunction of signals when SARS-CoV-2 is attached to its ACE2 receptor. Under this condition Ang(1-7) synthesis through ACE2 is likely decreased, Ang II accumulates and, increased Ang II binding to AT1R/AT2R leads to proinflammatory signals that ultimately will trigger both tissues damage (in particular lungs and heart) and hypertension. The lower-right panel illustrates that during COVID-19, the ACE2 mRNA expression is reduced leading to low surface expression of ACE2 (and plasma sACE2), AngII is in excess in plasma, yet Ang (1-7) remains synthesized through the NEP/TOP alternative pathway where neprilysin (NEP) and thimet oligopeptidase (TOP) convert AngI to Ang (1-7).

During the quantitative analysis of membrane ACE2 by flow cytometry, no decrease in expression of this molecule was observed on PBMCs and populations of T-cells, B-cells and CD16^+^ monocytic/dendritic cells from patients. This was surprising given that there is a decrease in the expression of the ACE2 transcript in PBMCs of COVID-19 patients (this paper, Figure 1B). Possibly, the downmodulation of ACE2 on PBMCs is not found due to differences in the sensitivity of qRT-PCR and flow cytometry techniques. In addition, the mean expression of ACE2 on the surface of PBMCs does not exceed 8-25%, which is perhaps too low to measure detectable variations at the protein level. Yet we found a significant decrease in membrane-bound ACE2 in a population of CD14^+^/HLADR^+^ monocytes (this paper, Figure1D), that corroborates with the down regulation of ACE2 transcription in adherent (monocytes) cells (this paper, Figure 1E). It is interesting to note that this conventional population of CD14^+^/HLADR^+^ monocytes increase in the PBMCs while the CD14^low^CD16^+^ inflammatory monocytes decrease as they leave the bloodstream and homing tissues (including the lungs) in symptomatic COVID-19 patients (71). Moreover, we found a decrease in the plasma levels of sACE2 in COVID-19 patients (this paper, Figure 2A). This reduced plasma level of sACE2 is likely to be a direct consequence of the reduction in the expression of the ACE2 mRNA leading to a lower biosynthesis of the ACE2 glycoprotein rather than a decrease in SARS-CoV-2-induced ACE2 shedding. Indeed, similarly to other Sarbecoviruses it is expected that the binding of the SARS-CoV-2 spike to cell-membrane anchored ACE2 trigger the release of sACE2. It was previously reported that the binding of a recombinant SARS-CoV spike protein to ACE2 positive-cells down regulated ACE2 expression through release of sACE2 (72,73). In a murine animal model it was observed that AngII-mediated decrease of ACE2 at the surface of cardiomyocyte was accompanied by the concomitant increase in cell-surface expression of the ADAM17 sheddase (74,75), suggesting a possible cleavage of membrane-bound ACE2 and shedding of sACE2. It was also previously reported that a recombinant SARS-CoV expressing the HCoV-NL63 spike, another human coronavirus using ACE2 for cell entry, trigger shedding of sACE2 (76). Interestingly, after a decrease in the plasma level of sACE2 in COVID-19 patients (during the acute phase of viral persistence), we found that this level returns to normal values when the virus is no longer detectable in healed individuals. This molecule could be considered a possible candidate for monitoring the evolution of the COVID-19 disease.

The fact that SARS-CoV-2 binds to the ACE2 receptor has been the source of intense debate among cardiologists regarding the risk of “liaisons dangereuses” (dangerous link) between the virus and its receptor and the management of patients who have hypertension (39,75,77). The medical management of hypertension involves the use of inhibitors of the renin-angiotensin-aldosterone system (RAAS), such as Angiotensin Converting Enzyme Inhibitors (ACEIs) and Angiotensin II Receptor Blockers (ARBs), considered to upregulate the cell surface expression of ACE2, with the possible adverse effects of increasing these patients’ susceptibility to SARS-CoV-2 (64,78). Nonetheless, a recent study reported that ACEIs reduced ACE2 expression in lungs (79). For hypertensive patients already infected with SARS-CoV-2, the question has also arisen of temporarily discontinuation of the treatment during COVID-19 disease to avoid viral over-replication and rapid cell-to-cell propagation of the virus. Conversely, it was also considered that discontinuing treatment could also worsen the general health status of patients and that maintaining treatment could have a favorable effect by acting as a vasodilator, antioxidant and anti-inflammatory through the action of Ang (1-7) on MAS-receptors. It is expected that when ACE2 expression is reduced, there is an accumulation of AngII that activates the AT1R and AT2R receptors and increases blood pressure through vasoconstriction. Here we found that the plasma concentration of AngI and AngII are significantly higher in the COVID-19 patients and patients healed of COVID-19 than in healthy volunteers (this paper Figures 2B, 2C). It might have seemed reasonable to hypothesize that if, in COVID-19 patients, ACE2 expression was decreased and serum AngII levels increased, the risk of hypertension could be increased. In the COVID-19 group, 7 out of 30 patients had a history of HT. We did not find a greater tendency to HT for patients who were not treated for HT before the diagnosis of their COVID-19 was established. In this group of patients, only two cases of HT (patient code: IHU_44 with blood pressure of 180/30 and hypokalemia at 3.45 mmol/L, and IHU_46 with blood pressure of 137/65 and a kalemia of 4.1 mmol/L; 2/23, 8.7%), were discovered at the time of their clinical examination. In the group of patients healed from COVID-19, only two (IHU-6 and IHU_27 patients) remained HT but they were already known for HT prior to the discovery of their COVID-19. It is worth noting that both AngI and AngII remained very high in plasma samples of patients even after the virus was apparently cleared from patients.

The decreased expression of ACE2 was consistent with the observed accumulation of AngII in COVID-19 patients. Therefore, it may be surprising to find no significant difference between the three groups studied with respect to the quantification of the Ang(1-7) plasma levels. In particular, we noted strong Ang(1-7) expression in three COVID-19 patients (patients code: IHU_24, IHU_8 and IHU_13). The IHU_24 patient, a woman in her late-40’s with HT, expressed the highest plasma level of AngII; patient IHU_8, a woman in her early-80’s with HT, expressed a very high plasma level of sACE2, while the patient IHU_13 was a man in his early-50’s with no known history of HT. The reduction in transcription of the ACE2 gene should result in a reduced capacity to cleave AngII to Ang (1-7). However, Ang (1-7) production did not collapsed in COVID-19 patients and tended to be slightly higher than in healthy volunteers. It cannot be excluded that ACE2 mRNA expression in PBMCs is valuable for this tissues only and that ACE2 may remain expressed on some cell types leading to cleavage of AngII that accumulates in excess COVID-19 patients to produce Ang (1-7) (80). We found (this paper, Figures 1, S1) that cell-surface expression of ACE2 can be differentially modulated in cellular populations. However, it is much more likely that an alternative pathway for the biosynthesis of Ang (1-7), independent of ACE2, can contribute to the biosynthesis of Ang(1-7). The comparison of intrarenal AngI, AngII and Ang(1-7) in wild type mice and tisACE^-/-^ mice lacking the ACE, revealed that the AngI and AngII levels were decreased by 80% in tisACE^-/-^ mice, whereas the Ang(1-7) levels were sustained (81). It was also reported that Ang(1-7) can be formed directly from AngI through a pathway that is independent of AngII production using the endopeptidases neprilysin (NEP), a 95 kDa membrane-anchored metalloendopeptidase located on the vascular surface of blood vessels and thimet oligopeptidase (TOP), a 80KDa soluble metalloendopeptidase, which both hydrolyzes Ang I decapeptide at the Pro-Phe bond to form Ang(1-7) (82-84). This suggests that in COVID-19 patients the balance between AngII and Ang(1-7) is more complex than previously assumed since the plasma levels of AngII are high but the levels of Ang(1-7) do not drop down. This should be further studied, as Ang(1-7) generation was previously reported capable to achieve protection against AngII-mediated lung injury and cardiovascular risks (77,85,86). However, the plasma levels of AngII in COVID-19 is probably too high so that its harmful effects are offset by the action of Ang(1-7). Moreover, it is likely that the increase in AngII mediate a dysregulation of the RAAS axis, with AngII-induced overproduction of aldosterone through AT1 receptor stimulation which may be at least in part responsible of the hypokaliemia observed in some COVID-19 patients (e.g. patient IHU_44), as recently reported elsewhere (87).

In conclusion, we have demonstrated here for the first time that during the COVID-19 disease, a decrease in ACE2 mRNA expression, ACE2 cell-surface protein and plasma level of sACE2, is observed. As a result, the plasma levels of AngII increase due to the absence of metabolism of AngII to Ang (1-7). However, this has no direct impact on the plasma levels of Ang(1-7) which remain stable or even increase slightly. This suggests that when the ACE2 pathway is less efficient or totally ineffective, Ang(1-7) is probably produced by metabolism of AngI to Ang (1-7), likely by the neprilysin and/or thimet oligopeptidase. The high plasma concentration of AngI in COVID-19 patients should favor this pathway. Although the plasma concentration of Ang(1-7) remains elevated in COVID-19 patients, it appears to be insufficient to prevent the harmful effects of AngII that accumulates in COVID-19. Finally we also report preliminary evidence that being on anti-HT treatment seems to be beneficial since, in the group of patients healed from COVID-19, those on anti-HT treatment were those who had the closest average ACE2 expression levels to the average ACE2 mRNA expression levels observed in healthy volunteers, suggesting a more rapid return to normal *ACE2* gene transcription.

## Data Availability

ll Data used and mentioned in the manuscript are available and can be consulted upon request.

## Acknowledgements

We are grateful to all the clinical, technical and paramedical staffs of the hospitalization units and laboratories for their expert assistance. We thank the students and our colleagues from the Mediterranean Infection IHU who volunteered to join the healthy control group. We thank Aissatou Bailo-Diallo, Orlane Soldani, Andriamiharimamy Rajaonison and Youssouf Sérémé for helpful technical discussions. We thank the Cookie Trad company for English editing.

## Authorship

IOO, CM, DR and CD contributed to the design of the study and conceived the manuscript. PB, LM, JLM and CD prepared the research protocol for ethical review by the French Participants Protection (CPP). IOO performed the *in vitro* experiments. CM, PB, JCL, PP, AS, MM and JCL took care of the COVID-19 patients. CM prepared Table I. IOO prepared the figures. CD supervised the work and wrote the paper. DR obtained the funding for this study. All authors reviewed and approved the final version of the manuscript

---

**Supplementary figure S1:**
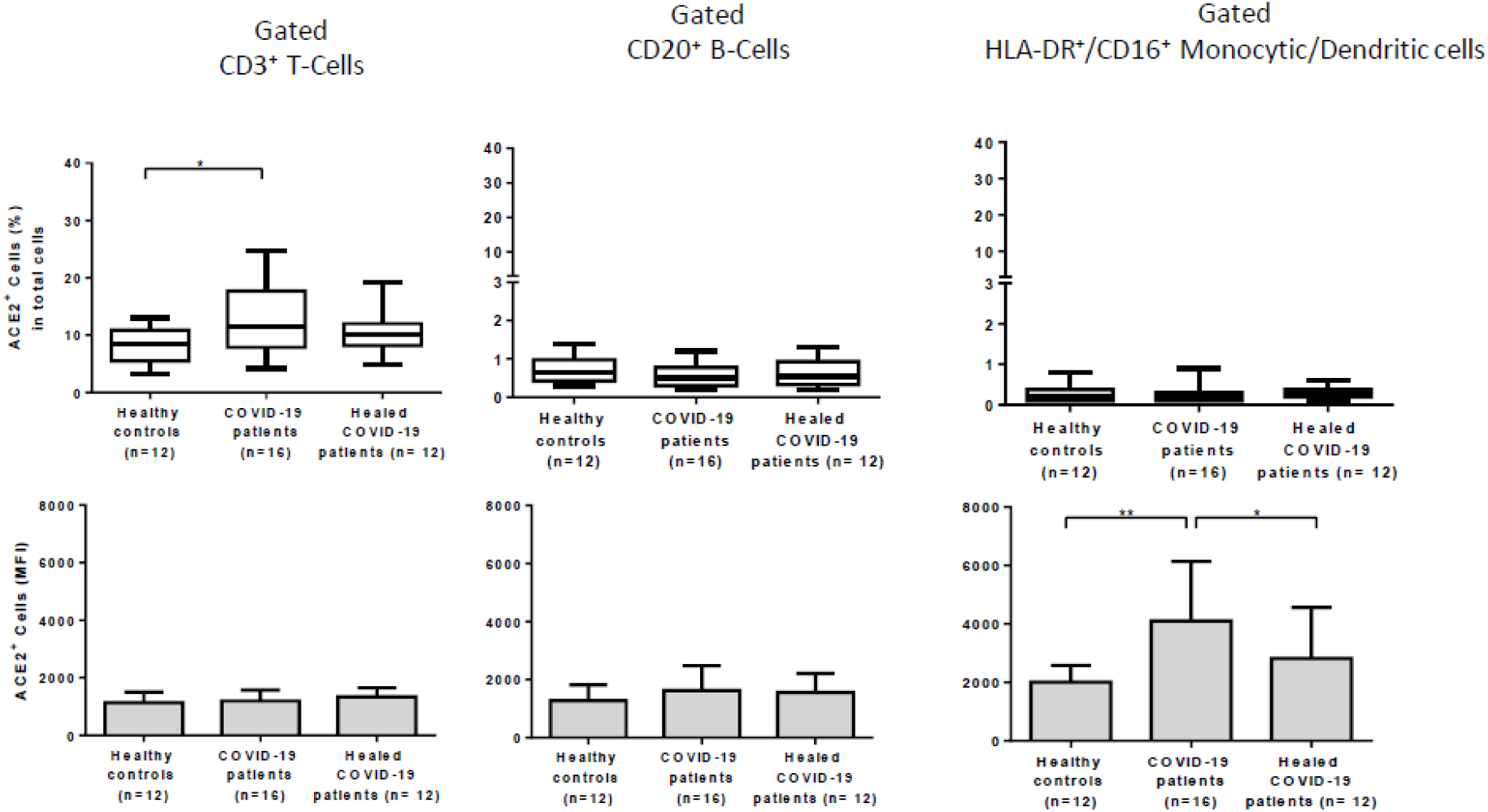
Histogram summarizing the data of flow cytometry analysis of ACE2 expression at the surface of T-cells, B-cells and monocytes. Comparison of percent cells expressing ACE2 and ACE2 MFI on cell samples from COVID-19 patients (n=12), patients healed from COVID-19 (n=12) and healthy volunteers (n=12). The gating was performed using different cluster differentiation-specific mAb allowing to select subpopulation of lymphocytes CD3^+^ T-cells, CD20^+^ B-cells and HLADR^+^/CD16^+^ monocytic/dendritic cells. The upper panels indicate the percent of cells expressing ACE2 with respect to the cells subpopulation analyzed while the lower panels are the histograms of ACE2 cell surface expression (MFI).

